# The PS4-Likelihood Ratio Calculator: Flexible allocation of evidence weighting for case-control data in variant classification

**DOI:** 10.1101/2024.04.09.24305536

**Authors:** Charlie F. Rowlands, Alice Garrett, Sophie Allen, Miranda Durkie, George J. Burghel, Rachel Robinson, Alison Callaway, Joanne Field, Bethan Frugtniet, Sheila Palmer-Smith, Jonathan Grant, Judith Pagan, Trudi McDevitt, Terri McVeigh, Helen Hanson, Nicola Whiffin, Michael Jones, Clare Turnbull, CanVIG-UK

## Abstract

**Background:** Within the 2015 American College of Medical Genetics/Association of Molecular Pathology (ACMG/AMP) variant classification framework, case-control observations can only be scored dichotomously as ‘strong’ evidence (PS4) towards pathogenicity or ‘nil’.

**Methods:** We developed the PS4-likelihood ratio calculator (PS4-LRCalc) for quantitative evidence assignment based on the observed variant frequencies in cases and controls. Binomial likelihoods are computed for two models, each defined by pre-specified odds ratio (OR) thresholds. Model one represents the hypothesis of association between variant and phenotype (e.g. OR≥5) and model two represents the hypothesis of non-association (e.g. OR≤1).

**Results:** PS4-LRCalc enables continuous quantitation of evidence for variant classification expressed as a likelihood ratio (LR), which can be log-converted into log LR (evidence points). Using PS4-LRCalc, observed data can be used to quantify evidence towards either pathogenicity or benignity. Variants can also be evaluated against models of different penetrance. The approach is applicable to balanced datasets generated for more common phenotypes and smaller datasets more typical in very rare disease variant evaluation.

**Conclusion:** PS4-LRCalc enables flexible evidence quantitation on a continuous scale for observed case-control data. The converted LR is amenable to incorporation into the now widely used 2018 updated Bayesian ACMG/AMP framework.

## Introduction

The American College of Medical Genetics/Association of Molecular Pathology (ACMG/AMP) published in 2015 a provisional framework for the interpretation and classification of genomic sequence variants^1^. Codes and weightings were provided for evidence items comprising the frequency of variant observations in humans with and without phenotype, predictions for sequence changes of protein and splicing impact and assays of variant function. The intention of the 2015 ACMG/AMP framework was to improve the consistency and robustness of variant classifications, with the authors providing prescriptive criteria and quantitative thresholds for many evidence items. Nevertheless, the authors recognised the 2015 framework to be provisional; a number of modifications and specifications to the original framework have been subsequently developed through international expert consensus. The ClinGen Sequence Variant Interpretation Group (SVI) has developed specifications for more general evidence items, whilst more specifications relating to individual genes or sets of genes have been developed by Variant Curation Expert Panels (VCEPs)^2-11^.

In the original 2015 ACMG/AMP framework, four ordinal evidence weightings were delineated (supporting, moderate, strong or very strong), with specification for how evidence items attaining these weightings were to be combined to provide overall classifications. In 2018, Tavtigian and SVI colleagues proposed a Bayesian reconfiguration of the framework, whereby evidence would instead be quantified as likelihood ratios (LRs, also termed OddsPath, odds of pathogenicity)^12^. By taking the logarithm (to base 2.08), these LRs might be translated into exponent (evidence) points (EPs), in such a way that the previous evidence weightings were transformed into a geometric progression of supporting (1 EP), moderate (2 EPs), strong (4 EPs) and very strong (8 EPs; Table 1)^13,14^. Assuming a prior probability of pathogenicity of 10%, EPs can be summed (or the product of the LRs calculated) and can then be converted to posterior probabilities and assigned to one of five overall variant classifications: benign (<0.1% probability of pathogenicity), likely benign (0.1-10%), variant of uncertain significance (VUS; 10-90%), likely pathogenic (90-99%) and pathogenic (>99%). The SVI and VCEPs have applied this LR-based approach to quantify the evidence weighting for data relating to functional assays (PS3/BS3), phenotype specificity (PP4), and in silico predictions (PP3/BP4)^15-19^. It has been advised the forthcoming revision of the 2015 ACMG/AMP framework will adopt the LR-EP system and that non-integer EPs may be permissible, a substantial evolution from the confined prescriptions for evidence combinations laid out in the original 2015 framework^20^.

**Table 1.** Flexible LR-based assignment of evidence weightings, as described in the 2018 Bayesian evolution of the ACMG/AMP framework. As described by Tavtigian et al., evidence criteria for which a likelihood ratio towards pathogenicity can be quantified may be converted to exponent points (EPs) through log-transformation (to base 2.08)^13,21,23^. This continuous approach reflects evidence strength quantitatively, in contrast to the categorical approach of the 2015 ACMG/AMP framework.

| Likelihood ratio range | Evidence points | Direction of evidence | ACMG/AMP (2015) Evidence strength |
| --- | --- | --- | --- |
| $\geq 350$ ($2.08^8$) | 8 | Towards pathogenicity | Very strong |
| $\geq 18.7$ ($2.08^4$) & $< 350$ | 4 | Towards pathogenicity | Strong |
| $\geq 4.33$ ($2.08^2$) & $< 18.7$ | 2 | Towards pathogenicity | Moderate |
| $\geq 2.08$ ($2.08^1$) & $< 4.33$ | 1 | Towards pathogenicity | Supporting |
| $\leq 0.48$ ($2.08^{-1}$) & $> 0.23$ | -1 | Towards benignity | Supporting |
| $\leq 0.23$ ($2.08^{-2}$) & $> 0.053$ | -2 | Towards benignity | Moderate |
| $\leq 0.53$ ($2.08^{-4}$) & $> 0.0029$ | -4 | Towards benignity | Strong |
| $\leq 0.0029$ ($2.08^{-8}$) | -8 | Towards benignity | Very strong |

One of the most fundamental observations indicating that a variant is disease-associated (i.e. pathogenic) is observation of that variant at a higher frequency in individuals with the relevant disease/phenotype (cases) than in those without (controls). Such case-control evidence was assigned code PS4 in the 2015 ACMG/AMP framework^1^. In the 2015 framework paper, discussion by authors highlighted that the strength of association might vary between different gene-phenotype dyads, as well as that the precision of the estimated effect size (i.e. the confidence interval) was an important consideration alongside the point estimate; the recommendation in the paper was allocation of ‘strong’ evidence for PS4 where OR>5 and the lower 95% CI>1^1^. However, PS4 is one of the few codes for which there has been no subsequent specification by the SVI, with the stipulations by VCEPs varying widely around how evidence might be allocated for PS4 ^7,11^. The overlap of use of the same datasets for PS4 as with codes for variant frequency in controls (PM2, BA1, BS1) and lack of provision for case-control data evaluation towards benignity have also been recognised as current limitations.

There is therefore requirement for a mechanism by which to translate across from the frequentist stipulations relating to case-control odds ratios (as per the current 2015 ACMG/AMP framework) into a Bayesian quantitation of a likelihood ratio (commensurate with the 2018 SVI framework evolution and forthcoming ACMG framework revision)^13,21^. Recently, Kanti et al. analysed SNP-array dataset on 75,657 breast cancer cases and 52,987 controls of European ancestry, calculating age-specific log-relative risk from survival analysis, to generate LRs for 24 *BRCA1* and 68 *BRCA2* variants^22^. Such methodology is well-suited for comprehensive prospective analyses of large case-control datasets where individual-level annotations for age and other parameters are available. However, we also require tools accessible to clinical diagnostic scientists to empower flexible and accurate quantitation of evidence from summary-level case-control data in the context of reactive classification of clinically identified variants.

We present here the PS4-likelihood ratio calculator (PS4-LRCalc; Figure 1), by which the observed frequency of a given variant in a series of cases can be compared to the observed frequency in a series of controls to quantitatively compare (i) the likelihood of the variant having an effect size at (or above) a specified level (target OR of association) to (ii) the likelihood of the variant having an effect size at (or below) a specified level (target OR of non-association). The ratio of these two likelihoods generates an LR that, when converted as described above, provides EPs of the form used within the Bayesian-points-ACMG framework, as described by Tavtigian et al.^12,23^. If derived from independently ascertained case-control series, these points can then be summed to generate a combined PS4 score.

**Figure 1.**
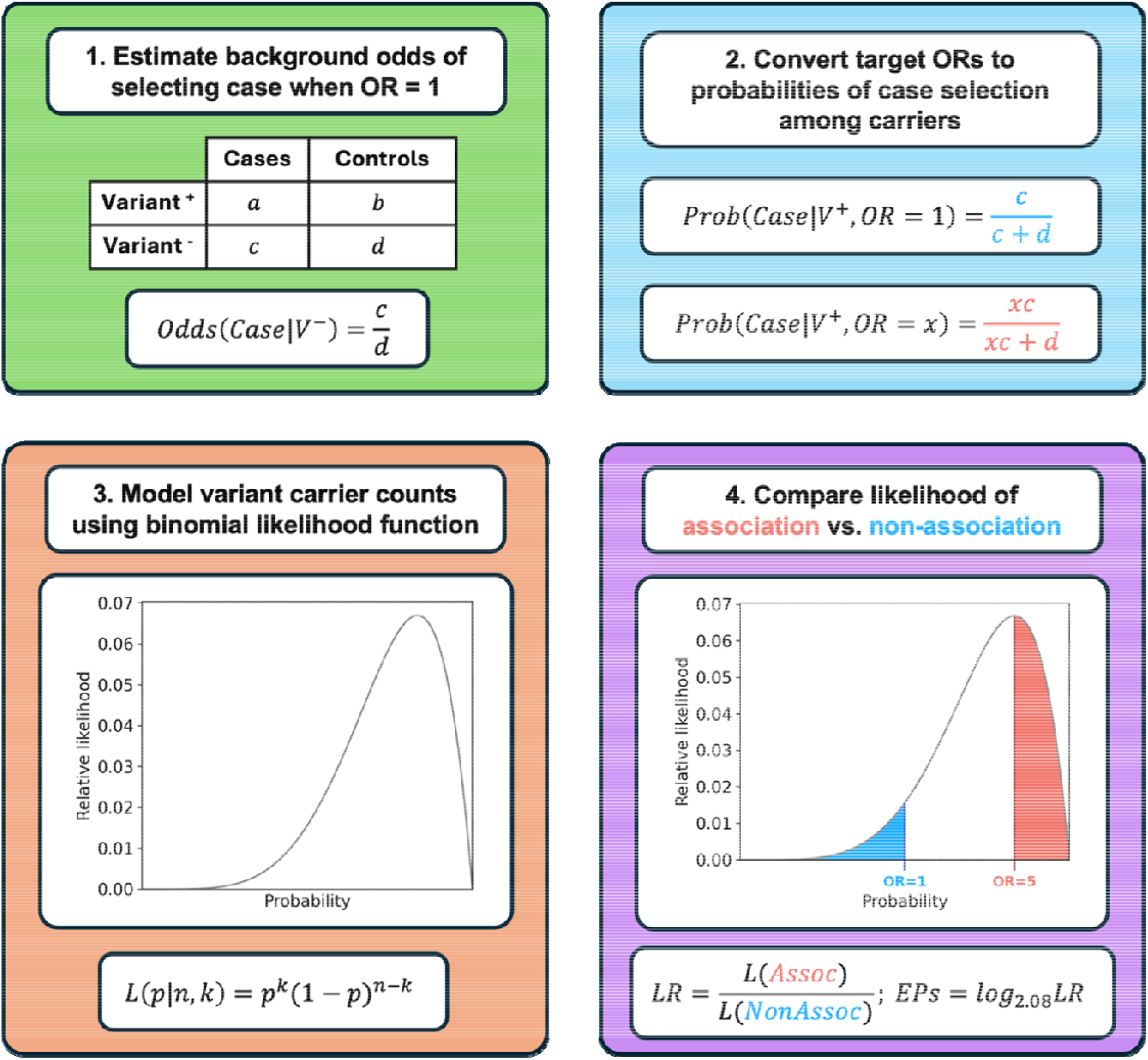
Overview of PS4-LRCalc framework for flexible PS4 application. (1) For a given set of case-control variant observations, the expected background odds of selecting a case among variant carriers are calculated using the equivalent observed odds among non-carriers. (2) The background odds are then scaled according to the ORs of association and non-association and converted to an expected probability of selecting a case among variant carriers under each hypothesis. (3) Variant observations in cases and controls are then modelled using a binomial likelihood function, which evaluates the likelihood that a given probability (p) of case selection would generate the observed data (k variant observations in cases across n total observations); note that these probabilities directly convert to odds values, which in turn generate a continuum of odds ratios across all possible values of p. (4) The likelihood ratio towards pathogenicity is determined by quantifying the total likelihood of association (L(Assoc); red area under curve) and dividing it by the total likelihood of non-association (L(NonAssoc); blue area under curve). The likelihood ratio (LR) can then be converted to Tavtigian exponent points (EPs) by taking its log (to base 2.08).

## Methods

### Derivation of likelihood ratio (LR)

We assume that observations of variant counts in cases and controls follow a binomial distribution. We use the binomial likelihood function to compute likelihoods for two models that represent competing hypotheses about the risk of disease associated with a specific variant. The first model, the hypothesis of association, is that the underlying effect size generating the observed variant distribution comprises an OR greater than the stated target OR of association, for example OR≥5. The second model, the hypothesis of non-association, is that the variant in not disease-causing; that is, the underlying effect size is less than the stated target OR of non-association, typically OR≤1 (noting that this may also encompass a protective effect; see Table 1). The likelihoods of the hypotheses of association and non-association, given the observed data, are hereafter termed the likelihood of association and likelihood of non-association, respectively (see Supplementary Methods for additional detail and worked example).

The PS4-likelihood ratio (PS4-LR) towards pathogenicity (equivalent to the odds of pathogenicity in the Bayesian framework described by Tavtigian et al.) is calculated by dividing the likelihood of association by the likelihood of non-association^13,21^. The LR is then converted to a logarithm (of base 2.08) to generate a log likelihood ratio (LLR), also termed exponent points or evidence points (EPs), which correspond to PS4 evidence weighting in the 2015 ACMG/AMP framework, as shown in Table 1.

PS4-LRCalc was developed in Python (v3.11), and analyses were performed using PyCharm (v23.1.1, Professional Edition) for remote development on a high-performance computing cluster. The online tool for LR calculator use was developed using Shiny for Python.

## Results

### Quantifying evidence towards pathogenicity: large, balanced case-control datasets (cancer susceptibility genetics scenario)

In Tables 2 and 3 and Figure 2a-f, we present illustrative scenarios of hypothetical variants observed in 10,000 cases and 10,000 controls, applying for our hypothesis of association a target OR of 5. This OR was selected for demonstration on account of being the threshold for disease association proposed in the 2015 ACMG/AMP framework. Thus, in each case a likelihood ratio (LR) is generated from comparison of the likelihood of the true underlying OR being ≥5 against the likelihood of the true underlying OR being ≤1 (the target OR of non-association). In scenarios 1-3 (Table 2), ‘strong’ evidence would have been awarded in the existing 2015-ACMG/AMP framework PS4 specification (2015-ACMG-PS4), as the observed OR exceeds 5 and lower 95% confidence interval exceeds one. However, the magnitude and confidence of association represented by these three scenarios of observed data vary widely: the LRs vary 10^11^-fold and EPs range from 6.1 to 43.0. Conversely, in scenarios 4-6 (Table 2), no evidence would be allocated under 2015-ACMG-PS4. However, in scenario 4, based on the observed data, the likelihood of association is more than five-fold greater than the likelihood of non-association: this equates to 2.3 EPs. In scenario 5, one fewer instance of the variant was observed in the case series compared to scenario 4, meaning the observed OR is lower (OR=4.0; 95% CI = 0.45-35.80) but nevertheless, the confidence interval readily encompasses the target OR of association (OR=5), and the likelihood of association (that the true underlying is OR≥5) is more than two-fold greater than the likelihood of non-association (that the true underlying OR≤1), translating to 1.2 EPs. Similarly, in scenario 6 (Figure 1c), the observed OR is 4.5 (0.97-20.84): whilst no evidence would be allocated under 2015-ACMG-PS4, the likelihood of association (OR≥5) is almost seventeen-fold greater than the likelihood of non-association (OR≤1), which equates to 3.9 EPs.

**Table 2.** Exemplar case-control scenarios generating exponent points (EPs) towards pathogenicity when using PS4-LRCalc. Shown are illustrative sets of variant observations in case and control datasets of equal size (10,000 individuals each) and the respective likelihood ratio (LR) and exponent points (EPs) generated for each using PS4-LRCalc. In scenarios 1-3, assignation of PS4 at ‘strong’ would have been possible under the 2015 ACMG/AMP framework; integration of PS4-LRCalc allows more refined quantification of evidence strength, such that the equivalent of ‘very strong’ evidence can be applied for scenarios 2 and 3. The variants depicted in scenarios 4-6 fail to fulfil the prescriptive 2015 ACMG/AMP criteria for application of PS4. Use of PS4-LRCalc, by contrast, allows application of PS4 at evidence strengths that are attenuated when compared to scenarios 1-3. Scenarios 1, 3 and 6 are further illustrated in Figure 2a-c.

| Scenario number | Cases | | Controls | | Observed effect size | | | | | ACMG 2015 framework (OR≥5, LCI≥1) | Likelihood of OR≥5 vs Likelihood of OR≤1 | |
| --- | --- | --- | --- | --- | --- | --- | --- | --- | --- | --- | --- | --- |
|  | Cases with variant | Total Cases | Controls with variant | Total Controls | Odds Ratio | Lower 95% CI | Upper 95% CI | P (chi-squared) | P (Fishers exact) |  | LR | EPs |
| 1 | 10 | 10,000 | 2 | 10,000 | 5.0 | 1.10 | 22.84 | 0.04 | 0.04 | STRONG | 33.28 | 4.8 |
| 2 | 100 | 10,000 | 20 | 10,000 | 5.0 | 3.12 | 8.15 | 4.71x10<sup>-13</sup> | 4.69x10<sup>-14</sup> | STRONG | 3.97x10<sup>13</sup> | 42.8 |
| 3 | 20 | 10,000 | 2 | 10,000 | 10.0 | 2.34 | 42.87 | 2.87x10<sup>-4</sup> | 1.20x10<sup>-4</sup> | STRONG | 2.35x10<sup>4</sup> | 13.7 |
| 4 | 5 | 10,000 | 1 | 10,000 | 5.0 | 0.58 | 42.82 | 0.22 | 0.22 | None | 5.29 | 2.3 |
| 5 | 4 | 10,000 | 1 | 10,000 | 4.0 | 0.45 | 35.80 | 0.37 | 0.37 | None | 2.41 | 1.2 |
| 6 | 9 | 10,000 | 2 | 10,000 | 4.5 | 0.97 | 20.85 | 0.07 | 0.07 | None | 16.79 | 3.9 |

**Table 3.** Exemplar case-control scenarios generating exponent points (EPs) towards benignity when using PS4-LRCalc. Shown are illustrative sets of case-control observations in datasets of equivalent size and their equivalent likelihood ratios (LRs) and exponent points (EPs) under the PS4-LRCalc model. The 2015 ACMG/AMP framework does not permit the use of lack of case-control signal as evidence for benignity. However, in scenarios 7-9, calculation of LRs using PS4-LRCalc allows application of PS4 in the benign direction at increasing strength – from the equivalent of moderate to very strong – as the number of variant observations increases. In scenarios 10 and 11, EPs for variants with ostensibly protective effects, i.e. observed at higher frequency in controls than cases, reach the equivalent of ‘very strong’ in the benign direction. Scenarios 8, 9 and 10 are further illustrated in Figure 2d-f.

| Scenario number | Cases |  | Controls |  | Observed effect size |  |  |  |  | ACMG 2015 framework (OR≥5, LCI≥1) | Likelihood of OR≥5 vs Likelihood of OR≤1 |  |
| --- | --- | --- | --- | --- | --- | --- | --- | --- | --- | --- | --- | --- |
|  | Cases with variant | Total Cases | Controls with variant | Total Controls | Odds Ratio | Lower 95% CI | Upper 95% CI | P (chi-squared) | P (Fishers exact) |  | Likelihood ratio | LLR: Exponent (evidence) points |
| 7 | 1 | 10,000 | 1 | 10,000 | 1.0 | 0.06 | 15.99 | 1.00 | 1.00 | None | 0.15 | -2.6 |
| 8 | 2 | 10,000 | 2 | 10,000 | 1.0 | 0.14 | 7.10 | 1.00 | 1.00 | None | 7.10x10<sup>-2</sup> | -3.6 |
| 9 | 10 | 10,000 | 10 | 10,000 | 1.0 | 0.42 | 2.40 | 1.00 | 1.00 | None | 3.75x10<sup>-4</sup> | -10.8 |
| 10 | 2 | 10,000 | 4 | 10,000 | 0.5 | 0.09 | 2.73 | 0.68 | 0.69 | None | 2.59x10<sup>-3</sup> | -8.1 |
| 11 | 2 | 10,000 | 10 | 10,000 | 0.2 | 0.04 | 0.91 | 0.04 | 0.04 | None | 1.55x10<sup>-7</sup> | -21.4 |

**Figure 2.**
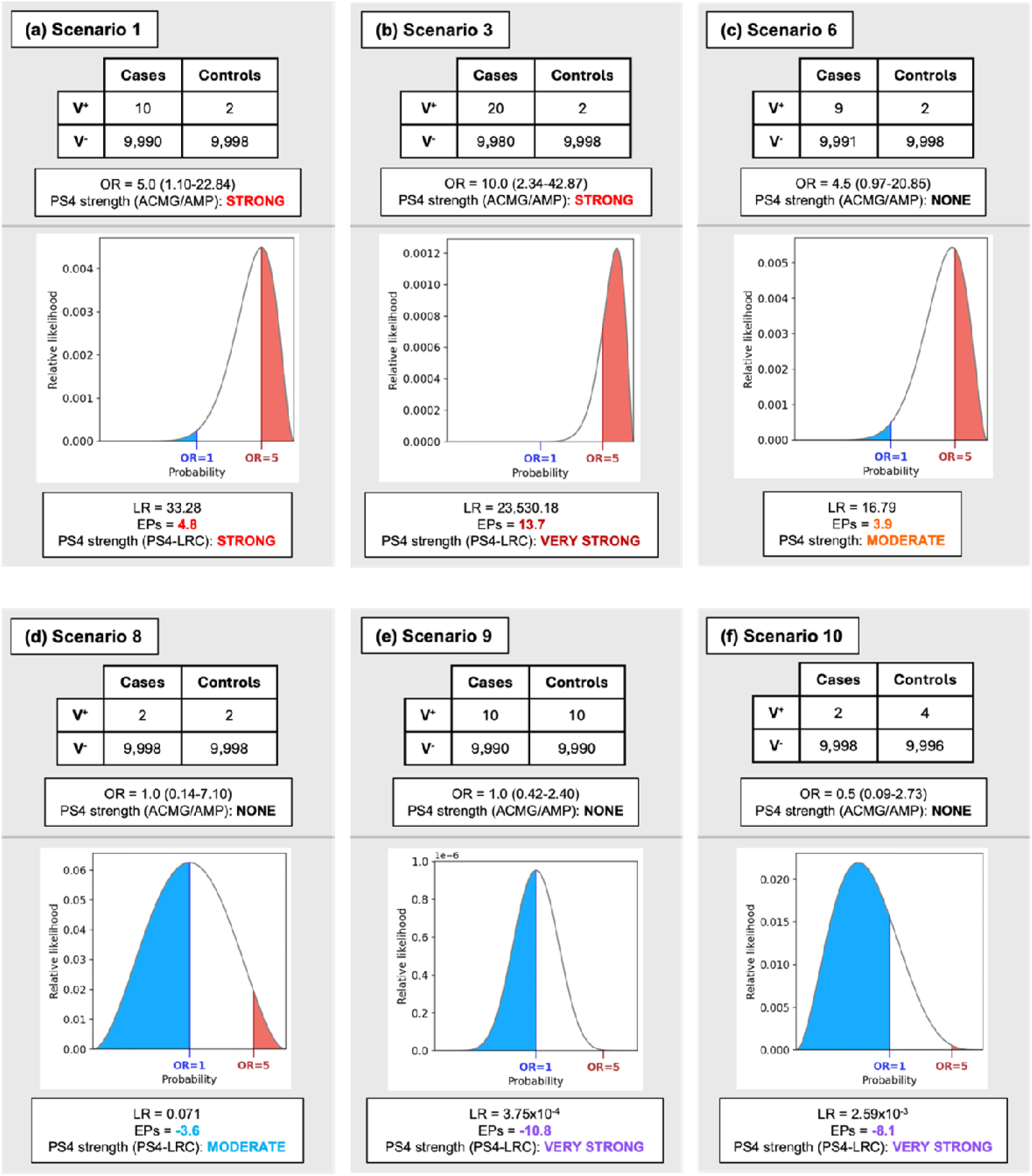
Comparison of applicable strength for the PS4 criterion between the 2015 ACMG/AMP guidelines and PS4-LRCalc for selected case-control scenarios. Counts of carriers (V^+^) and non-carriers (V^-^) of a variant between cases and controls are illustrated for exemplar scenarios indicative of (**a-c**) pathogenicity and (**d-f**) benignity shown in Tables 2 and 3. In the PS4-LRCalc approach, the distribution of variant carrier observations between cases and controls is modelled using a binomial likelihood curve, in which the likelihood of association (OR≥5, red) is divided by the likelihood of non-association (OR≤1, blue) to generate a likelihood ratio (LR) towards pathogenicity, that can then be converted to Tavtigian exponent points (EPs) towards pathogenicity or benignity and a corresponding applicable evidence strength under the 2015 ACMG/AMP framework. Notably, our approach allows assignation of EPs for scenarios that may not fulfil the existing ACMG/AMP PS4 guidance, including in support of benignity.

### Quantifying evidence towards benignity

PS4-LRCalc also enables quantitation of evidence towards benignity, as illustrated in Table 3. Again, we considered in each case the likelihood of association (target OR=5) versus the likelihood of non-association (target OR=1) based on observed data. In scenarios 7-9, we illustrate a range of scenarios in which the observed odds ratio is OR=1, but with increasing numbers of variant observations there is increasingly stronger evidence provided towards benignity. In scenarios 10 and 11, the frequency of the variant in controls is greater than that in cases, providing increasingly powerful evidence towards benignity.

### Quantifying evidence towards pathogenicity: different models of penetrance

In clinical cancer susceptibility genetics, genes associated with breast cancer are deemed to be of high penetrance if the association between pathogenic variants in that gene and phenotype is typically of OR≥4 (for example *BRCA1, BRCA2*), whilst genes for which pathogenic variants are typically of effect size (OR) 2-4 are deemed to be of moderate penetrance (for example, *CHEK2*). However, for some variants in *BRCA1* and *BRCA2*^24^, observed data suggest reduced penetrance for breast cancer of OR=2-4, risks more comparable to those ascribed to moderate penetrance genes^25^. Guidance exists for clinical management of patients with these reduced penetrance *BRCA1*/*BRCA2* variants. It may therefore be of utility on occasion to be able to assess *BRCA1*/*BRCA2* case-control variant data against both models of penetrance: considering firstly evidence of association at standard high penetrance (OR≥4) and for evidence of association at reduced penetrance (OR≥2). For example, if a variant is observed at a frequency of 12/10,000 in cases and 6/10,000 in controls, these observations would not constitute evidence for association against a target OR of association of 4, whilst against a target OR of 2 these observations would constitute moderate evidence (LR=5.5, EP=2.3; see Supplementary Table 1).

### Quantifying evidence towards pathogenicity: unbalanced datasets with small case series (rare disease genetics scenario)

We present in Supplementary Table 2 illustrative ultra-rare disease-type scenarios; that is, small numbers of variant observations in modest-sized case series being compared to large population control cohorts. We illustrate the impact of varying the hypothesis of association, considering OR≥1000, OR≥100 and OR≥10 in keeping with effect sizes commensurate with very rare Mendelian diseases.

### Approaches to accommodate data uncertainty

The PS4-LRCalc approach provides, based on the observed data, quantitation of the comparative likelihoods of the underlying OR being at or above a higher value (target OR of association) versus at or below another, lower value (target OR of non-association). PS4-LRCalc will inherently reflect sample size (power, sampling variability), namely that the magnitude of the LR attained for a variant of a given frequency and strength of disease association will be determined by the magnitude of the case and control data series. If the observed data are accurate and robust, then this LR most directly quantifies the evidence towards pathogenicity (or benignity) afforded from the observed data.

However, on occasion there may be uncertainty regarding the accuracy of genotyping or phenotyping of the case (and/or control) series. This is an issue inherent to any application of case (or control) data towards variant classification, rather than the issue being particular to the PS4-LRCalc. However, by virtue of its parameterisation, PS4-LRCalc affords various options for introducing ‘conservatism’ to the case-control analysis, the selection and specification of which will be predicated on the level of uncertainty of data accuracy and/or desire for conservatism:

i. **Adjustment of the opposing hypothesis:** quantitation of evidence can be rendered more conservative by adjusting the competing hypothesis. That is, where unadjusted data provides evidence towards pathogenicity, the target OR of non-association may be increased. In Supplementary Table 3, we illustrate how the LR/LLR are impacted by comparing a target OR of association of OR=5 to different target ORs of non-association, namely OR=1, OR=2 and OR=5.
ii. **Sensitivity analysis:** by reducing the number of case observations and/or increasing control observations (commensurate with degree of uncertainty and ‘trust’ in the data), it is possible to assess the robustness of the unadjusted case-control signal. For example, if the count of case observations with the variant is n=2, reducing the case count to n=1 and reconducting LR quantification may inform the confidence around the original prediction. The logical extension of this principle is that the n=1 case variant scenario should be sensitivity-tested at n=0 (i.e. therefore n=1 case series are effectively disallowed). This strategy may be particularly pertinent in rare disease scenarios in which the case denominator is modest, meaning that each observation of an instance of the variant in a case is contributing substantially to the evidence weighting.
iii. **Application of a confidence interval to the target ORs** (the target OR of association and the target OR of non-association): rather than using the target OR for association (e.g. OR=5), it is possible to utilise the lower (70%, 90% or 95%) confidence interval of this estimate, as derived from the expected counts under the hypothesis of association. Rather than using the target OR for non-association (e.g. OR=1), it is possible to utilise the upper (70%, 90% or 95%) confidence interval of this estimate, as derived from the expected counts under the hypothesis of non-association (see Supplementary Methods). An LR incorporating one or both of these values is thus attenuated (i.e. more conservative) compared to an LR derived using the direct target ORs. Of note, where there are low total variant observations and/or unbalanced case-control dataset size this will have a substantive impact on the standard error of the OR, such that addition of a confidence interval will result in accordingly aggressive diminution of the LR (i.e. this approach may be highly punitive in these scenarios, Supplementary Table 4).

There is an additional rules-based restriction which may warrant consideration to avoid generation of an LR from comparison of two hypotheses each of miniscule likelihood. In some scenarios with well-powered case-control signal, the target ORs of non-association and association may lie below and above the confidence interval limits of the observed OR estimate, respectively. In these scenarios, the likelihoods of association and non-association will constitute only a minuscule proportion of the total likelihood space. However, if the miniscule likelihood of the observed data hypothesis of association is substantially greater than the miniscule likelihood for the hypothesis of non-association, an LR of sizeable magnitude can be generated (Supplementary Figure 1). In such a scenario in practice, there is high confidence of an effect intermediate between the two hypotheses, meaning that interrogation against a much higher target OR of association is thus unlikely to be a clinically meaningful endeavour.

### Accessible tool for direct access to PS4-LR-calculator tool

An online Shiny for Python tool is available at https://turnbull-lab.shinyapps.io/ps4_lrcalc/. This allows the input of (i) case and control variant observations and denominators, (ii) a target odds ratio of association, (iii) a target odds ratio of non-association, and (iv) (optional) confidence intervals. The outputs include the relative likelihoods, a likelihood ratio, evidence points and a distribution curve of likelihoods.

## Discussion

We present PS4-LRCalc, which enables quantification of evidence towards variant pathogenicity/benignity based on a continuous output from a statistical model rather using a dichotomous evidence allocation. The PS4-LRCalc approach allows evaluation of the observed data against a pre-specified ‘target OR of association’ and a pre-specified ‘target OR of non-association’, thus quantified as an LR which effectively serves to bridge the frequentist-Bayesian divide. This approach is applicable across a spectrum of use-cases, including both the highly penetrant effects observed from small case series in investigation of ultra-rare Mendelian disease, as well as less penetrant effects inferred from the larger case series available for the investigation of more common phenotypes for example for variants in cancer susceptibility genes. The advantages of the PS4-LRCalc approach include:

- Firstly, the evidence is quantified as a LR, which is then converted into an LLR, which equates to a number of exponent (evidence) points. This approach is consistent with the 2018 Tavtigian-SVI adaptation of the 2015 ACMG framework which is to be adopted in the forthcoming ACMG framework update.
- Secondly, the evidence strength (LLR) is quantified on a continuous scale, affording direct quantitative reflection of the magnitude of evidence towards pathogenicity afforded by the observed data. This offers dramatically improved flexibility compared to the 2015-ACMG-PS4 specification of dichotomous options of strong evidence or no evidence.
- Thirdly, the parameterisation of the PS4-LRCalc model allows specification of target ORs of interest. This enables flexibility around specifying these hypotheses, for example, examining models of different penetrance.
- Fourthly, this approach allows ready combination of evidence from multiple (independent) studies/sources, with summing of evidence points (or multiplication of likelihood ratios).
- Fifthly, this approach allows quantitation of evidence towards benignity based on observed data; arguably an elegant complement or alternative to the current BA1/BS1 codes, by which evidence towards benignity is assessed based purely on variant frequency in controls.
- Finally, we outline several potential approaches by which to manage uncertainty inherent to clinical data of uncertain quality/provenance.

Application of summary-level case-control data for variant interpretation carries a number of cautions and caveats: these apply equally to the existing 2015-ACMG-PS4 as to the PS4-LRCalc approach:

- There should be appropriate matching of ethnicity between case and control series.
- The standards for sequencing/genotyping and downstream QC (quality control) must be considered, in particular where they differ between cases and controls.
- The provenance of the data (for example, if extracted from a publication) is particularly important where variant numbers are small (meaning that single observations may substantially influence the outcome).
- If there is any level of enrichment or over-selection amongst the case series, this will cause inflation in the observed OR in relation to that of an unselected cohort. If the cohorts have been genotyped for a specific variant of known effect size, an ‘enrichment factor’ for the cohort could be calculated, by which observed ORs may be suitably down-adjusted. Finally, using summary-level variant frequencies in cases and controls will disregard differential variant distribution in the context of variable age-related penetrance (namely, where the variant is disproportionately frequent in younger age groups, in which background rates of disease are lower).

We have created an accessible methodology and user-friendly publicly available tool enabling flexible, accurate quantitation for variant classification of case-control data as a likelihood ratio and respective exponent evidence points. In particular, this approach affords (i) allocation of lower levels of contributary evidence in instances for which the confidence and/or effect size would not attain ‘strong’ by the dichotomous cut-off of 2015-ACMG-PS4 (ii) allocation of higher than ‘strong’ levels of contributary evidence where supported by observed data, and (iii) evidence towards benignity. We anticipate this type of approach and tool will be of utility to diagnostic clinical scientists and clinicians with increased availability of newly collated large diagnostic testing and population sequencing datasets, in particular on update of the ACMG/AMP framework, in which evidence quantitation will be more continuous and utilise an LR/LLR framework.

## Data Availability

All data produced in the present work are contained in the manuscript

## Acknowledgement

We would like to acknowledge the Institute of Cancer Research-Royal Marsden Biomedical Research Centre. We would like to acknowledge the contribution of Chey Loveday to initial development of the methodology.

## Author Contributions

Conceptualization: C.T.; Data curation: C.F.R.; Formal analysis: C.F.R.; Funding acquisition: C.T.; Investigation: C.F.R.; Methodology: C.T., M.J., N.W., A.G., S.A., C.F.R.; Project administration: S.A.; Software: C.F.R.; Supervision: C.T.; Visualization: A.G., S.A., C.F.R.; Validation: M.D., G.J.B., A.C., J.F., B.F., S.P-S., J.G., T.McD., T.McV., H.H., N.W. Writing-original draft: C.T., A.G., S.A., C.F.R.; Writing-review & editing: all authors.

