## Supplementary Material for "The PS4-Likelihood Ratio Calculator: Flexible allocation of evidence weighting for case-control data in variant classification"

### Supplementary Materials

#### Supplementary Methods

*Derivation of likelihood ratio (LR)*

In order to calculate a likelihood ratio towards pathogenicity for observed case-control data of the type illustrated below:

|  | **Cases** | **Controls** |
| --- | --- | --- |
| **Variant^+^** | 5 | 1 |
| **Variant^-^** | 9995 | 9999 |
| **Total** | 10000 | 10000 |

We first use the expected effect size of pathogenic variants in the gene of interest to define a target odds ratio (OR) for our hypothesis of association (i.e. towards pathogenicity), for example OR≥5, and our hypothesis of non-association (i.e. towards benignity), typically OR≤1. A graphical representation of this and subsequent steps are shown in Figure 1.

The upper ($p_{u}$) and lower ($p_{l}$) thresholds of probability for each of our hypotheses are then calculated, i.e. the probabilities of selecting a case from among all variant carriers that are expected to generate the range of odds pertaining to each hypothesis. By default, we define $p_{l}$ for the hypothesis of non-association as 0 (as decreasing odds tend towards a probability of 0) and the $p_{u}$ for the hypothesis of association as 1 (as increasing odds tend towards a probability of 1). The thresholds relating to the lower boundary of the hypothesis for association and upper boundary of the hypothesis of non-association are calculated using the following formula:

$$p=\frac{xc}{xc+d}$$

Where:

- $x$ is the threshold odds ratio for the hypothesis under consideration (in this exemplar, 5 for the hypothesis of association and 1 for the hypothesis of non-association)
- $c$ is the number of variant non-carriers in the case series
- $d$ is the number of variant non-carriers in the control series
- NB: when $x=1$, $p = \frac{c}{c+d}$, which is equal to the background probability of selecting a case from all variant non-carriers

We then use the $p$ values derived from the odds of association and non-association as the lower and upper boundaries for the hypotheses of association and non-association, respectively.

In the exemplar scenario above, the lower and upper thresholds of probability for the hypothesis of association (OR≥5) are calculated as:

$p_{l} =\frac{5\times9995}{5\times9995+9999}= 0.833$and $p_{u} = 1$

While for the hypothesis of non-association (OR≤1), these values are:

$p_{l} = 0$ and $p_{u}=\frac{9995}{9995+9999}\approx0.5$

Observations of variants in cases and controls are then modelled using a binomial likelihood function. This describes the likelihood that any given probability of success, $p$, is true given our observed data ($n,k$):

$$L\left( p;n,k \right)=p^{k}\times\left( 1-p \right)^{\left( n-k \right)}$$

Where:

- $n$, the number of trials, is the sum of cases and controls with a variant
- $k$, the number of successes, is the number of cases with the variant
- $p$, the probability of success, is expressed as a range of values separately for each hypothesis.

In the context of PS4-LRCalc, this function generates a curve representing the likelihood that a given probability of selecting a case among variant carriers would give rise to the observed data. The summed likelihood of each hypothesis is thus the area under the binomial likelihood curve bounded by the upper and lower probability thresholds of that hypothesis.

The summed likelihoods for each hypothesis are calculated using numerical integration via Gaussian adaptive quadrature:

$$L_{H}=\int_{p_{l}}^{p_{u}} f\left( x \right) dx$$

Where:

- $p_{l}$ is the probability value defining the lower boundary of the hypothesis under consideration.
- $p_{u}$ is the probability value defining the upper boundary of the hypothesis under consideration.
- $f\left( x \right)$ is the probability density function of the binomial likelihood.
- dx represents an infinitesimally small change in the variable $x$, indicating integration with respect to $x$.

#### The ratio of the summed likelihoods is then derived as follows:

$$LR=\frac{L_{H1}}{L_{H0}}$$

Where:

$L_{H1}$ is the summed likelihood for the hypothesis of association

$L_{H0}$ is the summed likelihood for the hypothesis of non-association

The value of this ratio indicates how likely the hypotheses are relative to one another. A value greater than 1 indicates that the hypothesis of association is more likely than the hypothesis of non-association, given the observed data, and vice versa. In the exemplar, division of the summed likelihood yields a likelihood ratio (LR) of 5.29, which is positive and therefore indicates an increased likelihood of pathogenicity compared to benignity.

Finally, the likelihood ratio is converted to a log likelihood ratio (LLR) by calculating the log to base 2.08. The LLR is also termed evidence points or exponent points (EPs):

$$EPs={log}_{2.08}LR$$

Positive exponent values are considered as evidence towards pathogenicity, whereas negative values are evidence towards benignity. In the exemplar, the LR of 5.29 obtained through division of summed likelihoods is equivalent to 2.27 EPs.

*Calculation of adjusted odds thresholds using expected counts*

As described in the main text, certain clinical scenarios may necessitate incorporation of conservatism into LR estimates. In the PS4-LRCalc model, the probability thresholds of association and non-association correspond to the odds estimate that is most likely to yield the respective target OR. We have devised a methodology for introduction of conservatism that moves the probability threshold up (for the hypothesis of non-association) and/or down (for the hypothesis of association) to account for the fact that, for a given set of observed data, odds values at some distance from the estimated odds threshold may still yield a an OR for which the confidence interval encompasses the target OR. The distance of this movement is reflective of the overall levels of confidence afforded by the specific distribution of observed counts.

For a set of variant and non-variant observations in a case-control scenario, a contingency table of observed data can be derived as follows:

|  | **Cases** | **Controls** |
| --- | --- | --- |
| **Variant+** | $a$ | $b$ |
| **Variant-** | $c$ | $d$ |

Where the odds ratio is ordinarily calculated as:

$$OR= \frac{\frac{a}{b}}{\frac{c}{d}}$$

To generate confidence intervals around the odds thresholds reflecting the hypotheses of association and non-association, we first construct a contingency table reflecting the expected values of $a$ and $b$, assuming $c$ and $d$ to be constant, under each hypothesis. It can be shown algebraically that the updated tables with expected counts $a'$ and $b'$ can be constructed as follows:

|  | **Cases** | **Controls** |
| --- | --- | --- |
| **Variant+** | $a' =\frac{xc(a+b)}{xc+d}$ | $b' =\frac{d(a+b)}{xc+d}$ |
| **Variant-** | $c$ | $d$ |

Where $x$ = the target OR and $a$-$d$ are the original observed values, as above.

The confidence interval around the OR estimate generated by the expected counts is then calculated as standard for a contingency table:

$$CI=e^{ln(\frac{\frac{a'}{b'}}{\frac{c}{d}})\pm x\sqrt{\frac{1}{a'}+\frac{1}{b'}+\frac{1}{c}+\frac{1}{d}}}$$

Where $x$ is the number of standard errors expected to generate the desired level of confidence under a normal distribution (1.96 for 95%; 1.645 for 90% etc.). For the hypothesis of non-association, the upper CI limit of the expected counts is returned as the updated odds threshold; for the hypothesis of association, the lower limit is returned.

Supplementary Figures

**Supplementary Figure 1.** *Illustration of a high-confidence odds ratio that is intermediate between competing hypotheses of association and non-association.* Shown is the contingency table and corresponding binomial likelihood curve for a variant count distribution representing an odds ratio (OR) of 2.02 (95% CI: 1.59-2.57). The high confidence of this OR means the point estimate and 95% CI lie entirely between the two competing target ORs of non-association and association (1 and 5, respectively). This means the areas under the curve corresponding the likelihood of association and non-association occupy only the tails of the binomial curve and so do not meaningfully convey the likelihood distribution, which is centred far outside the two considered hypotheses. In such situations, the point estimate of the OR is already well-estimated and so implementation of PS4-LRCalc will not be of clinical utility.

|  | **Case** | **Control** |
| --- | --- | --- |
| **Variant^+^** | 200 | 100 |
| **Variant^-^** | 9800 | 9900 |

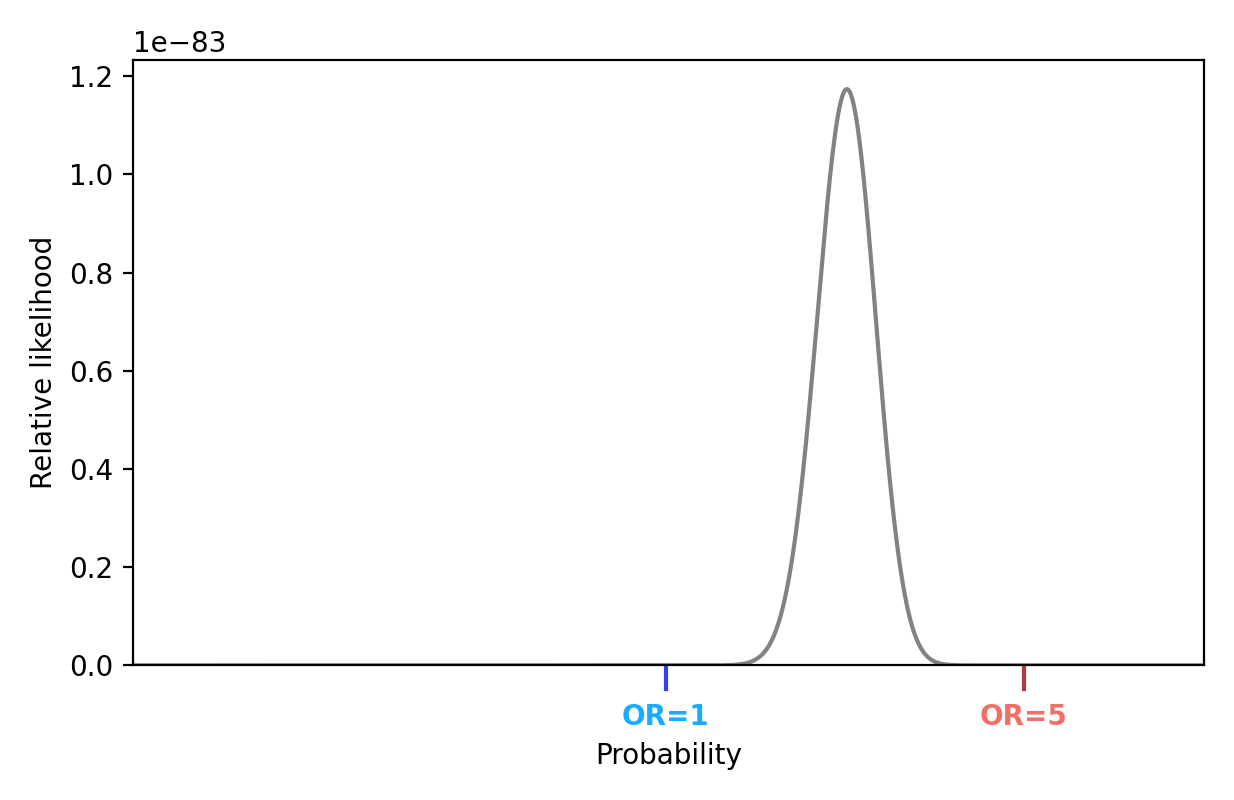

#### Supplementary Tables

Below are depicted extended scenarios investigating the impact of parameter variability on the output of the PS4-LRCalc model. In each case the specified numbers of variant observations in cases and controls are presented alongside their respective likelihood ratio (LRs) and exponent points (EPs) under particular parameter combinations.

**Supplementary Table 1.** *Impact of decreasing target OR of association (hypothesis of reduced penetrance)*. Shown are three scenarios which fail to attain PS4 under the 2015 ACMG/AMP specification. PS4-LRCalc allows awarding of PS4 at the equivalent of supporting strength for scenario 12 when applying a hypothesis of association of OR≥4 (a common standard for high penetrance in breast cancer susceptibility genes (BCSGs)). On reducing the hypothesis of association to OR≥2 (a widely used threshold for intermediate-penetrance BCSGs), all three variants attain evidence for PS4. PS4-LRCalc thus facilitates the examination of individual variants under multiple models of variant effect, thereby permitting consideration of intermediate penetrance.

|  | **Cases** | | **Controls** | | **Observed effect size** | | | | | **ACMG 2015 framework (OR≥4, LCI≥1)** | **Likelihood OR≥4 vs. OR≤1** | | **ACMG 2015 framework (OR≥2, LCI≥1)** | **Likelihood OR≥2 vs. OR≤1** | |
| --- | --- | --- | --- | --- | --- | --- | --- | --- | --- | --- | --- | --- | --- | --- | --- |
| **Scenario #** | **# w/ var** | **Total** | **# w/ var** | **Total** | **OR** | **Lower 95% CI** | **Upper 95% CI** | **p-value (χ^2^)** | **p-value (Fisher exact)** |  | **LR** | **EPs** |  | **LR** | **EPs** |
| 12 | 6 | 10,000 | 2 | 10,000 | 3.0 | 0.61 | 14.87 | 0.29 | 0.29 | NONE | 2.92 | 1.5 | NONE | 6.94 | 2.7 |
| 13 | 3 | 10,000 | 1 | 10,000 | 3.0 | 0.31 | 28.85 | 0.62 | 0.62 | NONE | 1.40 | 0.5 | NONE | 2.88 | 1.4 |
| 14 | 12 | 10,000 | 6 | 10,000 | 2.0 | 0.75 | 5.33 | 0.24 | 0.24 | NONE | 0.81 | -0.3 | NONE | 5.49 | 2.3 |

**Supplementary Table 2.** *Simulation of rare disease-type scenarios.* In the six scenarios shown here, the frequency of variants in small-to-modest case series is evaluated against a population-scale dataset, as may be typical in a rare disease clinical setting. All scenarios would attain PS4 under the 2015 ACMG/AMP criterion; application of PS4-LRCalc under hypotheses of association of OR≥10 and OR≥100 suggests applicability of varying numbers of EPs equivalent to “very strong” for all scenarios. At a higher target OR of association of 1000, scenario 18, with an observed OR of ~50 instead is ascribed EPs towards benignity. These use cases assume fully robust genotyping and phenotyping: sensitivity assessment and adjustment may be warranted in scenarios where numbers are small and there is uncertainty regarding the robustness of data quality/ascertainment.

|  | **Cases** | | **Controls** | | **Observed effect size** | | | | | **ACMG 2015 framework (OR≥5, LCI≥1)** | **Likelihood OR≥10 vs. OR≤1** | | **Likelihood OR≥100 vs. OR≤1** | | **Likelihood OR≥1000 vs. OR≤1** | |
| --- | --- | --- | --- | --- | --- | --- | --- | --- | --- | --- | --- | --- | --- | --- | --- | --- |
| **Scenario #** | **# w/ var** | **Total** | **# w/ var** | **Total** | **OR** | **Lower 95% CI** | **Upper 95% CI** | **p-value (χ^2^)** | **p-value (Fisher exact)** |  | **LR** | **EPs** | **LR** | **EPs** | **LR** | **EPs** |
| 15 | 1 | 30 | 2 | 300,000 | 5,172.4 | 456.32 | 58,628.79 | 4.62x10^-183^ | 3.00x10^-4^ | STRONG | 1.78x10^7^ | 22.8 | 1.78x10^7^ | 22.8 | 1.71x10^7^ | 22.7 |
| 16 | 2 | 30 | 2 | 300,000 | 10,700.0 | 1457.78 | 78,746.26 | 0.00 | 5.80x10^-8^ | STRONG | 1.23x10^11^ | 34.9 | 1.23x10^11^ | 34.9 | 1.22x10^11^ | 34.9 |
| 17 | 5 | 30 | 20 | 300,000 | 2,999.8 | 1044.01 | 8,619.48 | 0.00 | 3.73x10^-16^ | STRONG | 1.30x10^19^ | 60.1 | 1.30x10^19^ | 60.1 | 1.28x10^19^ | 60.1 |
| 18 | 1 | 300 | 20 | 300,000 | 50.2 | 6.71 | 374.98 | 9.35x10^-4^ | 2.00x10^-2^ | STRONG | 4.34x10^3^ | 11.4 | 1,746.75 | 10.2 | 0.03 | -5.0 |
| 19 | 2 | 300 | 20 | 300,000 | 100.7 | 23.43 | 432.58 | 1.96x10^-23^ | 2.27x10^-4^ | STRONG | 5.86x10^5^ | 18.1 | 3.84x10^5^ | 17.6 | 20.63 | 4.1 |
| 20 | 5 | 300 | 20 | 300,000 | 254.2 | 94.78 | 681.85 | 1.39x10^-176^ | 5.03x10^-11^ | STRONG | 4.91x10^12^ | 39.9 | 4.79x10^12^ | 39.9 | 7.03x10^9^ | 31.0 |

**Supplementary Table 3.** *Scenarios investigating the impact of increasing the hypothesis of non-association.* The six scenarios from Table 1 (main manuscript) were each evaluated against alternative hypotheses of non-association (OR≤2 and OR≤5), to illustrate one potential approach for introduction of conservatism to LR estimates. Increasing the hypothesis of non-association is commensurately punitive to the outputted signal of pathogenicity, with only scenario 23 (of observed OR 10, markedly higher than the target OR of association) remaining eligible for application of PS4 towards pathogenicity.

|  | **Cases** | | **Controls** | | **Observed effect size** | | | | | **ACMG 2015 framework (OR≥5, LCI≥1)** | **Likelihood OR≥5 vs. OR≤1** | | **Likelihood OR≥5 vs. OR≤2** | | **Likelihood OR≥5 vs. OR≤5** | |
| --- | --- | --- | --- | --- | --- | --- | --- | --- | --- | --- | --- | --- | --- | --- | --- | --- |
| **Scenario #** | **# w/ var** | **Total** | **# w/ var** | **Total** | **OR** | **Lower 95% CI** | **Upper 95% CI** | **p-value (χ^2^)** | **p-value (Fisher exact)** |  | **LR** | **EPs** | **LR** | **EPs** | **LR** | **EPs** |
| 21 | 10 | 10,000 | 2 | 10,000 | 5.0 | 1.10 | 22.84 | 0.04 | 0.04 | STRONG | 33.28 | 4.8 | 2.69 | 1.4 | 0.59 | -0.7 |
| 22 | 100 | 10,000 | 20 | 10,000 | 5.0 | 3.12 | 8.15 | 4.71x10^-13^ | 4.69x10^-14^ | STRONG | 3.97x10^13^ | 42.8 | 20,832.96 | 13.6 | 0.89 | -0.2 |
| 23 | 20 | 10,000 | 2 | 10,000 | 10.0 | 2.34 | 42.87 | 2.87x10^-4^ | 1.20x10^-4^ | STRONG | 23,530.18 | 13.7 | 114.34 | 6.5 | 3.23 | 1.6 |
| 24 | 5 | 10,000 | 1 | 10,000 | 5.0 | 0.58 | 42.82 | 0.22 | 0.22 | NONE | 5.29 | 2.3 | 1.26 | 0.3 | 0.49 | -0.96 |
| 25 | 4 | 10,000 | 1 | 10,000 | 4.0 | 0.45 | 35.80 | 0.37 | 0.37 | NONE | 2.41 | 1.2 | 0.75 | -0.4 | 0.36 | -1.4 |
| 26 | 9 | 10,000 | 2 | 10,000 | 4.5 | 0.97 | 20.84 | 0.07 | 0.07 | NONE | 16.79 | 3.9 | 1.8 | 0.8 | 0.48 | -1.0 |

**Supplementary Table 4.** *Introduction of conservatism through addition of confidence intervals*. The introduction of confidence intervals to target OR thresholds (as detailed in Supplementary Methods) may allow introduction of conservatism. The six scenarios from Table 1 (main manuscript) were each evaluated with the introduction of confidence intervals of various magnitude – 70%, 90% and 95% – around the target ORs of association (lower confidence interval) and non-association (upper confidence interval). While the incorporation of 70% CIs leads to relatively modest decreases in the number of EPs applicable towards pathogenicity, 90% and 95% CIs are more punitive, with more substantial attenuation of the original outputted LR signal.

|  | **Cases** | | **Controls** | | **Observed effect size** | | | | | **ACMG 2015 framework (OR≥5, LCI≥1)** | **Likelihood OR≥5 vs. OR≤1** | | | | | | | |
| --- | --- | --- | --- | --- | --- | --- | --- | --- | --- | --- | --- | --- | --- | --- | --- | --- | --- | --- |
|  |  |  |  |  |  |  |  |  |  |  | **No CI** | | **70% CI** | | **90% CI** | | **95% CI** | |
| **Scenario #** | **# w/ var** | **Total** | **# w/ var** | **Total** | **OR** | **Lower 95% CI** | **Upper 95% CI** | **p-value (χ^2^)** | **p-value (Fisher exact)** |  | **LR** | **EPs** | **LR** | **EPs** | **LR** | **EPs** | **LR** | **EPs** |
| 27 | 10 | 10,000 | 2 | 10,000 | 5.0 | 1.10 | 22.84 | 0.04 | 0.04 | STRONG | 33.28 | 4.8 | 7.66 | 2.8 | 3.79 | 1.8 | 2.8 | 1.4 |
| 28 | 100 | 10,000 | 20 | 10,000 | 5.0 | 3.12 | 8.15 | 4.71x10^-13^ | 4.69x10^-14^ | STRONG | 3.97x10^13^ | 42.8 | 5.10x10^10^ | 33.7 | 1.36x10^9^ | 28.7 | 2.3x10^8^ | 26.3 |
| 29 | 20 | 10,000 | 2 | 10,000 | 10.0 | 2.34 | 42.87 | 2.87x10^-4^ | 1.20x10^-4^ | STRONG | 23,530.18 | 13.7 | 737.18 | 9.0 | 141.57 | 6.8 | 67.98 | 5.8 |
| 30 | 5 | 10,000 | 1 | 10,000 | 5.0 | 0.58 | 42.82 | 0.22 | 0.22 | NONE | 5.29 | 2.3 | 2.49 | 1.2 | 1.73 | 0.7 | 1.48 | 0.5 |
| 31 | 4 | 10,000 | 1 | 10,000 | 4.0 | 0.45 | 35.80 | 0.37 | 0.37 | NONE | 2.41 | 1.2 | 1.71 | 0.7 | 1.39 | 0.4 | 1.26 | 0.3 |
| 32 | 9 | 10,000 | 2 | 10,000 | 4.5 | 0.97 | 20.84 | 0.07 | 0.07 | NONE | 16.79 | 3.9 | 5.1 | 2.2 | 2.87 | 1.4 | 2.23 | 1.1 |
